# Implementation strategy modifications: An applied multi-site comparison using ERIC and FRAME-IS for the “Fluoroquinolone Restriction for the Prevention of *Clostridioides difficile* infection Trial” (FIRST)

**DOI:** 10.64898/2026.04.28.26351921

**Authors:** Vishala Parmasad, Demetrius Solomon, Douglas Wiegmann, Marin Schweizer-Looby, Nasia Safdar

## Abstract

**Background:** Implementation strategies are dynamic techniques used to apply evidence-based practices (EBPs) to diverse contexts. Despite their importance, context-specific selection and modification of implementation strategies remain underreported, limiting understanding of how to optimize strategy deployment across heterogeneous healthcare settings. We describe a systematic method to document and analyze modifications to implementation strategies using four diverse hospital sites from the Fluoroquinolone Restriction for the Prevention of *Clostridioides difficile* infection (FIRST) trial as case studies.

**Methods:** FIRST was a multisite fluoroquinolone pre-prescription restriction intervention delivered via the electronic health record. We partnered with multidisciplinary stakeholders at each site to co-design and adapt the intervention using pre-planned implementation strategies. Multiple data sources (interviews, meeting notes, implementation diaries) collected iteratively over two years were analyzed to identify strategy modifications. Strategies were coded using Expert Recommendations for Implementing Change (ERIC) conceptual clusters, and modifications were documented using the Framework for Reporting Adaptations and Modifications to Evidence-Based Implementation Strategies (FRAME-IS). Modified strategies were categorized as planned or unplanned and contextualized via thematic content analysis.

**Results:** Across 458 total modifications, the most modified strategies focused on facilitating stakeholder engagement, adapting to local contexts, and using evaluative approaches to improve EBP uptake/sustainment. Planned modifications (n=330, 72%) outnumbered unplanned modifications (n=157, 34%). Rural and community hospitals required more unplanned modifications (average 41 vs. 31 for academic centers), while sites with prior restrictive intervention experience had higher planned-to-unplanned ratios (3.1:1 vs. 1.6:1). Academic hospitals with trainee rotations required ongoing education and higher strategy modifications. All modifications maintained EBP core fidelity. Site-specific patterns organizational characteristics were linked to modification intensity and type, including absorptive capacity, prior experience, relational coordination, rurality, and educational requirements.

**Conclusions:** Integrating ERIC and FRAME-IS enabled systematic documentation of implementation strategy modifications across diverse settings. Planned:unplanned modification ratios provided novel insights into organizational absorptive capacity and implementation preparedness. Standardized implementation approaches inadequately address critical organizational differences, requiring context-sensitive strategy selection and intensity calibration. This work advances implementation science methodology by demonstrating how systematic modification documentation can inform tailored implementation support.

**Trial Registration:** ClinicalTrials.gov Identifier: NCT03848689

## 1.0 Introduction

Increasing evidence from implementation science demonstrates that evidence-based practices (EBP) require contextually-relevant implementation strategies to fit with populations and service systems, and to produce effective implementation outcomes (adoption, fidelity, penetration, sustainability).(1–5) Proctor et al. (2013) define implementation strategies as “methods or techniques used to enhance the adoption, implementation, and sustainability of a clinical program or practice.”(6) Evidence supports multifaceted, multilevel implementation approaches combining and modifying discrete strategies for complex interventions.(7) However, insufficient implementation strategy documentation, reporting, and analysis limits systematic evaluation of strategy effectiveness, leading implementation researchers to describe implementation strategies as a “black box.”(2, 6, 8, 9) To address the gap in strategy documentation, Powell et al.’s (2015) Expert Recommendations for Implementing Change (ERIC) project used expert consensus via modified Delphi process to establish a taxonomy of 73 discrete implementation strategies with standardized definitions.(1) Waltz et al. (2015) identified nine conceptual strategy clusters into which ERIC strategies fall.(10) Widespread reference to these conceptual clusters provides opportunities for applied comparative assessment.(10, 11)

Research on implementation strategy modifications is growing; however gaps remain in standardized strategy and modification documentation, obscuring optimal modification-to-EBP matching and anticipation of context-driven adaptations.(12) Miller et al. (2021) developed the Framework for Reporting Adaptations and Modifications to Evidence-Based Implementation Strategies (FRAME-IS) to systematically document modifications.(13) FRAME-IS extends the original FRAME model by Stirman, Baumann and Miller (2019),(14) enabling structured documentation of who modified the strategies, what was modified, when, at what level, and whether modifications were planned or unplanned. FRAME-IS applied to multi-site analyses of antibiotic stewardship remains underexplored, despite increasing usage for a variety of intervention settings.(15–17)

To address this gap, we integrated ERIC strategy cluster definitions with FRAME-IS to document and analyze modifications made to implementation strategies in the Fluoroquinolone Restriction for the Prevention of *Clostridioides difficile* infection (FIRST) trial, a hybrid type-2 implementation-effectiveness trial.(18) Hybrid type-2 trials have coprimary aims to simultaneously test the efficacy of a clinical intervention and an implementation intervention or strategy.(18, 19) The goal of FIRST was to simultaneously implement and assess the effectiveness of an antibiotic stewardship effort to decrease fluoroquinolone (FQ) broad-spectrum antibiotic class prescribing. These antibiotics demonstrate a high association with hospital-acquired *Clostridioides difficile* infection (CDI) rates, which represent a significant public health burden.(20–22) Antibiotic prescribing restriction through pre-prescription authorization (PPA) has demonstrated efficacy in reducing CDI incidence.(23) However, implementing pre-prescription authorization interventions requires navigating complex organizational dynamics, prescriber behaviors, and workflow integration, exemplifying the challenges of translating evidence into varied contextual settings.

This study leverages FIRST trial data from four participating sites to demonstrate systematic integration of ERIC and FRAME-IS frameworks for documenting and analyzing implementation strategy modifications across heterogenous organizational contexts. We present a replicable method for documenting implementation strategy modifications and demonstrate how modification patterns relate to organizational characteristics, providing actionable insights for tailoring implementation support.

## 2.0 Materials and Methods

### 2.1 Study Design and Setting

The FIRST trial was an Agency for Healthcare Research and Quality (AHRQ)-funded multisite type II hybrid implementation-effectiveness trial.(18) It introduced an EHR-integrated FQ restriction intervention in intensive care units (ICUs) that met the inclusion criteria (including an existing antibiotic stewardship program without FQ restrictions, infectious disease and pharmacist support, EPIC EHR, EHR data extraction capacity). All sites were recruited after IRB approval and institutional leadership commitment and had dedicated on-site implementation coordinators and a facilitated implementation design.

### 2.2 Intervention Description

The FIRST intervention EHR best practice alert (BPA) was triggered when ICU prescribers ordered FQ antibiotics (e.g., ciprofloxacin, levofloxacin, moxifloxacin). The BPA: (a) allowed FQ prescribing for approved indications, (b) provided non-FQ alternatives for non-approved indications with rationale for substitution, and/or (c) required authorization from infectious disease physicians or antimicrobial stewardship pharmacists when providers wished to continue FQ prescribing for non-approved indications.

Approved indications were determined through stakeholder (i.e., infectious disease physician, infectious disease pharmacist, pharmacist, antibiotic stewardship team member, and intensivist) feedback at each site. Alternative antibiotic suggestions were embedded in the alert, providing immediate decision support. Authorization processes varied by site based on stewardship team availability: 24/7 pharmacist coverage (Sites A, B), business hours with on-call backup (Sites C, D), or attending physician authorization pathways.

### 2.3 Implementation Strategy Selection and Stakeholder Engagement

Initial implementation strategies were selected through structured meetings with multidisciplinary members of the research team at the EBP-originating pilot site, including infectious disease physicians, pharmacists, antimicrobial stewardship program leaders, information technologists, and clinical department chiefs. Core implementation strategies were pre-specified based on implementation science literature and pilot site experiences including stakeholder engagement meetings, facilitation through dedicated site implementation coordinators, educational sessions for prescribers, audit and feedback on FQ prescribing patterns, tailoring intervention components to local workflows, and iterative refinement based on formative evaluation data.

Stakeholder engagement occurred through regular meetings (weekly during intensive pre-implementation, bi-weekly during implementation, monthly during maintenance). Meeting agendas addressed implementation progress, barrier identification, strategy modification discussions, and action item assignment. All stakeholders had authority to propose modifications, with decisions made collaboratively.

### 2.4 Data Collection

Multiple data sources were collected at each site over 24 months (January 2020-December 2021). Data collection emphasized capturing (1) work system processes and (2) barriers and facilitators to the implementation during pre-best-practice alert (intervention) launch and post-intervention launch periods. Real-time documentation minimized retrospective recall bias.

- Implementation diaries (semi-structured surveys) of pre-launch activities designed by the implementation scientist and human factors engineers were completed by the site implementation coordinators (n=4) who included infectious disease pharmacists, infectious disease physicians, and a health researcher (public health MSc). These documented daily implementation activities, modification decisions, and contextual observations, as well as the time required.
- Facilitated implementation meeting notes (n=58 total across sites: Site A n=15, Site B n=22, Site C n=13, Site D n=8). Notes from iterative facilitation meetings between the study implementation lead (MBBS/ PhD in medical anthropology) and the site implementation coordinators were coded for strategy discussions, modification proposals, decision-making processes, and contextual factors influencing implementation.
- Site context questionnaires completed by the site implementation coordinators documenting: (1) ICU characteristics (e.g., no. of beds, open/closed, medical/surgical ICU, pharmacist staffing); (2) infection and control practices (e.g., infection preventionist staffing, educational efforts, surveillance and screening, etc.) and (3) antibiotic stewardship practices (staffing, facility-specific recommendations, stewardship culture-related questions).
- Semi-structured interviews conducted via videoconference 6 months post-intervention launch with key informants (n=-19 total across sites). Interviews were conducted by the study implementation lead and human factors engineers with: medical doctors (8), nurse (1), pharmacists (7), site implementation coordinators (4). Interview domains: implementation experiences, barrier encounters, facilitator identification, modification rationales, organizational context factors. Interviews were audio-recorded and transcribed. occurred at mid-implementation and post-implementation timepoints.
- Data collection emphasized capturing both planned strategy selections and unplanned modifications arising during implementation. Real-time documentation minimized retrospective recall bias.

### 2.5 Analysis Framework

Two researchers from different disciplines (systems engineering and medical anthropology/medical training) independently coded all data using MaxQDA qualitative analysis software. Coding proceeded iteratively with regular meetings to compare coding, discuss discrepancies, and achieve consensus. Statements fitting multiple categories were coded multiply.

Implementation strategies were systematically coded using ERIC’s 73 discrete strategies organized into nine conceptual clusters in accordance with Miller et al. (2021): (1) Use evaluative and iterative strategies; (2) Provide interactive assistance; (3) Adapt and tailor to context; (4) Develop stakeholder interrelationships; (5) Train and educate stakeholders; (6) Support clinicians; (7) Engage consumers; (8) Change infrastructure; (9) Utilize financial strategies.

Each modification was documented using FRAME-IS modules capturing seven domains:

- Module 1 (Description): EBP description, implementation strategy description, modification outline
- Module 2 (What): Modification type (content, context, training, evaluation)
- Module 3 (Nature): Specific modification characteristics (tailoring, adding, removing, lengthening, etc.) and fidelity implications
- Module 4 (Goal/Rationale): Why modification was made, at what level (organizational, implementer, clinician, patient)
- Module 5 (When): Timing (pre-implementation, implementation, maintenance) and planned vs. unplanned categorization
- Module 6 (Who): Participants in modification decision-making (program leaders, implementers, researchers)
- Module 7 (Spread): Scope of modification (organization-wide, clinic/unit level, individual level)

Planned vs. unplanned categorization criteria: Planned modifications were defined as those anticipated during pre-implementation planning, documented in meeting notes or other feedback before intervention launch, or identified as contingency plans. Unplanned modifications were reactive responses to unanticipated barriers, unexpected stakeholder resistance, unforeseen technical challenges, or external disruptions not planned for during pre-implementation. A third category, “planned-reactive,” captured anticipated potential issues requiring reactive implementation (e.g., knowing resident rotations would require ongoing education but not scheduling specific sessions in advance).

Rigor was maintained through multiple approaches: member checking with site implementation teams to verify interpretations, triangulation across multiple data sources, detailed documentation of analytic decisions by the interdisciplinary research team, and systematic tracking of data sources for all findings.

### 2.6 Contextual Analysis

Site-specific organizational characteristics were systematically documented through structured assessment tools completed by the site implementation coordinators: organizational size (bed capacity, ICU beds, annual admissions), academic status (teaching vs. non-teaching, trainee presence), geographic setting (urban vs. rural, regional referral patterns), prior restriction experience (presence/absence of existing antibiotic restrictions, antibiotic stewardship program details), stewardship infrastructure (dedicated pharmacist FTE, physician champions, data systems), IT capacity (EHR platform, build expertise, technical support availability), workforce composition (hospitalists vs. private attendings, resident training programs).

Thematic content analysis identified relationships between organizational characteristics and modification patterns. Analysis proceeded iteratively, with themes emerging from data and refined through team discussion. Cross-site comparisons highlighted patterns in modification types, quantities, and timing across different organizational contexts.

## 3.0 Results

### 3.1 Overview of Implementation Strategy Modifications

Across four sites (Table 1), 487 implementation strategy modifications were documented over 24 months. Planned modifications (n=330, 68%) substantially outnumbered unplanned modifications (n=157, 32%).

**Table 1:**
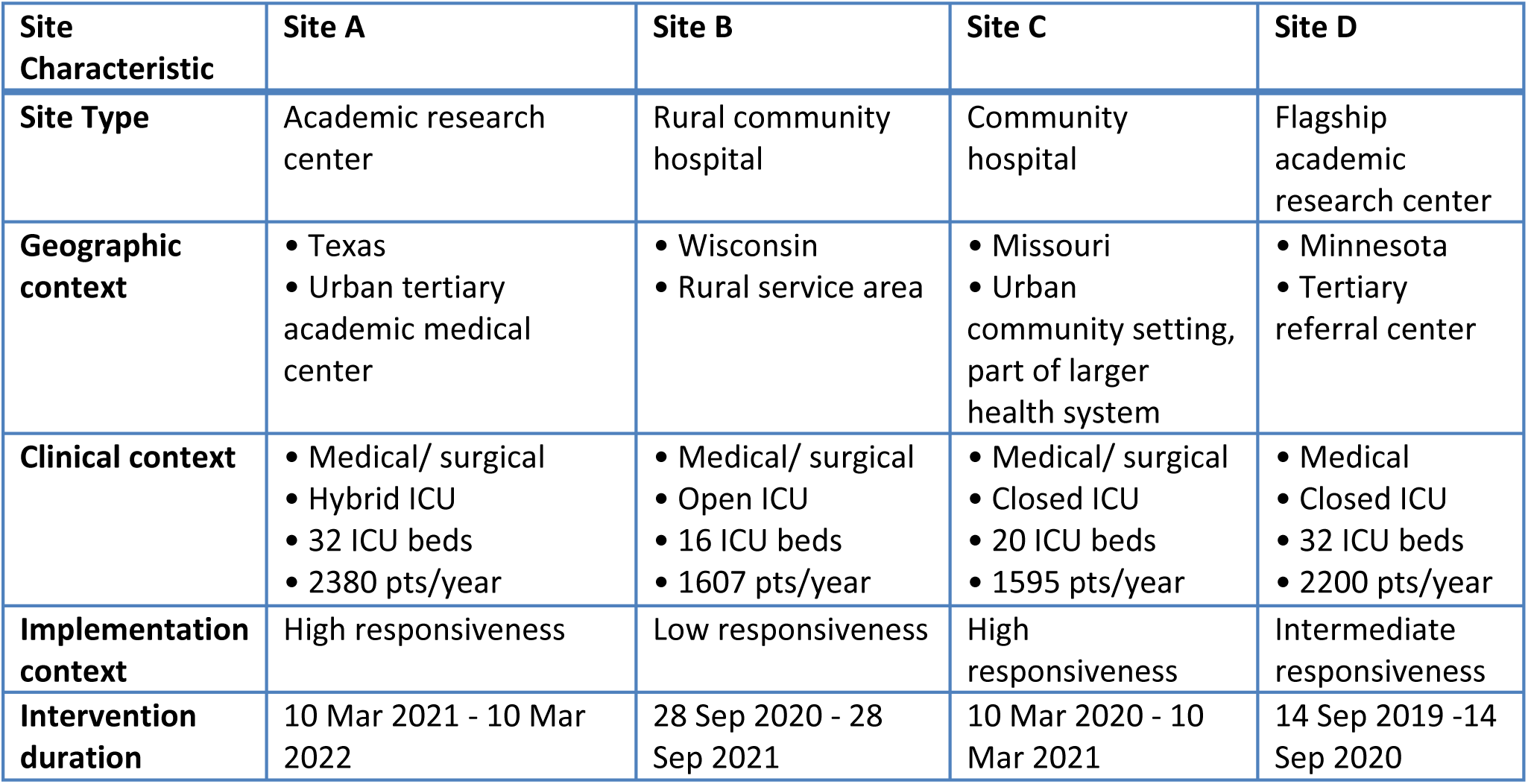
FIRST trial site characteristics.

| Site Characteristic | Site A | Site B | Site C | Site D |
| --- | --- | --- | --- | --- |
| Site Type | Academic research center | Rural community hospital | Community hospital | Flagship academic research center |
| Geographic context | • Texas<br>• Urban tertiary academic medical center | • Wisconsin<br>• Rural service area | • Missouri<br>• Urban community setting, part of larger health system | • Minnesota<br>• Tertiary referral center |
| Clinical context | • Medical/ surgical<br>• Hybrid ICU<br>• 32 ICU beds<br>• 2380 pts/year | • Medical/ surgical<br>• Open ICU<br>• 16 ICU beds<br>• 1607 pts/year | • Medical/ surgical<br>• Closed ICU<br>• 20 ICU beds<br>• 1595 pts/year | • Medical<br>• Closed ICU<br>• 32 ICU beds<br>• 2200 pts/year |
| Implementation context | High responsiveness | Low responsiveness | High responsiveness | Intermediate responsiveness |
| Intervention duration | 10 Mar 2021 - 10 Mar 2022 | 28 Sep 2020 - 28 Sep 2021 | 10 Mar 2020 - 10 Mar 2021 | 14 Sep 2019 -14 Sep 2020 |

Site-specific modification totals revealed substantial variation: Site A (large academic center with established restrictions) generated 126 total modifications (90 planned, 26 unplanned, 3.5:1 ratio); Site B (rural community hospital) generated 174 modifications (116 planned, 58 unplanned, 2.3:1 ratio); Site C (community hospital with stewardship) generated 94 modifications (58 planned, 37 unplanned, 1.6:1 ratio); Site D (academic center with limited BPA culture) generated 103 modifications (58 planned, 32 unplanned, 1.8:1 ratio).

The range in total modifications (94 to 174) and planned:unplanned ratios (1.6:1 to 3.5:1) suggested substantial contextual influence on both modification intensity and anticipatory planning capacity. Higher modification counts did not necessarily indicate implementation difficulty but rather reflected contextual complexity, stakeholder engagement intensity, and iterative refinement approaches.

### 3.2 Most Modified ERIC Strategy Clusters

Four ERIC clusters dominated modifications across all sites: (1) Train and educate stakeholders (n=88, 18.1%), (2) Adapt and tailor to context (n=86, 17.6%), and (3) Develop stakeholder interrelationships and Use evaluative and iterative strategies (n=79, 16.2% respectively). “Engage consumers” generated 58 modifications (11.9%). “Provide interactive assistance, “Change infrastructure,” “Support clinicians,” and “Utilize financial strategies” clusters accounted for remaining modifications (Table 2).

**Table 2:** ERIC Implementation Strategy Modifications Across Four Sites.

| Site Characteristic | Site A | Site B | Site C | Site D |
| --- | --- | --- | --- | --- |
| **TOTAL MODIFICATIONS** | 116 | 174 | 94 | 103 |
| **Planned (Proactive)** | 90 (78%) | 116 (67%) | 58 (62%) | 58 (64%) |
| **Unplanned (Reactive)** | 26 (22%) | 58 (33%) | 36 (38%) | 32 (36%) |
| **Planned:Unplanned Ratio** | 3.5:1 | 2.0:1 | 1.6:1 | 1.8:1 |
| **1st Most Modified** | Train/Educate (30) | Adapt/Tailor (38) | Use Evaluative (23) | Use Evaluative (21) |
| **2nd Most Modified** | Use Evaluative (20) | Develop Stakeholder (33) | Adapt/Tailor (19) | Develop Stakeholder (19) |
| **3rd Most Modified** | Develop Stakeholder (17) | Train/Educate (28) | Change Infrastructure (13) | Train/Educate (19) |
| **Most Unplanned Mods** | Train/Educate (10) | Adapt/Tailor (20) | Use Evaluative (14) | Train/Educate (10) |
| **No Unplanned Mods** | Support Clinicians, Provide Interactive, Change Infrastructure, Financial Strategies | Engaging Consumers, Change Infrastructure | Engaging Consumers, Develop Stakeholder, Financial Strategies | Change Infrastructure, Financial Strategies |
| **Pre-Implementation Phase** | Rapid (<6 months) | Extensive (12+ months) | Moderate (6-8 months) | Moderate (6-8 months) |
| **Implementation Scope** | ICU-focused (MICU, SICU) | ICU-focused (2 units) | Hospital-wide | ICU-only (MICU) |
| **Key Success Factors** | Existing restrictions, handshake rounds, CMO support | Multidisciplinary team, 24/7 ACS pharmacist | Established stewardship, skilled IT pharmacist | Data monitoring, senior physician engagement |
| **Unique Challenges** | Resident rotations, hybrid ICU | Resource constraints, rural workforce, COVID suspension | Private ID group, transfer workflow issues | BPA resistance, recent Epic transition |
| **COVID-19 Disruption** | Minimal - virtual grand rounds | Severe - 6-month suspension | Minimal - 1-month extension | Moderate - IT staff shortages |

Training and education modifications concentrated in Sites A and D (academic centers with rotating residents or trainees), requiring perpetual onboarding cycles. Modifications addressed timing (orientation sessions, monthly refreshers, just-in-time training), modality (in-person vs. virtual, individual vs. group), content tailoring (specialty-specific examples, role-based training), and reinforcement mechanisms (email reminders, reminders from lecturers, embedded EHR resources).

Adapt and tailor to context modifications focused on integration strategies that tailored the implementation to site workflows.

Stakeholder interrelationship development modifications emphasized building trust, establishing communication channels, addressing concerns, and maintaining engagement throughout implementation. Modifications included additional stakeholder meetings beyond protocol, expanded participation to previously uninvolved groups (e.g., surgical departments, nursing councils), creation of feedback loops, and establishment of ongoing communication mechanisms (newsletters, email updates, shared dashboards).

Evaluative strategy modifications reflected commitment to data-driven refinement. Modifications included enhanced data collection (expanding metrics beyond FQ use to alternative antibiotic patterns, resistance trends, clinical outcomes), increased reporting frequency (monthly to weekly during intensive implementation), tailored feedback formats (individual provider reports, departmental summaries, leadership dashboards), and responsive strategy adjustments based on evaluation findings.

### 3.3 Site-Specific Modification Patterns

#### Site A (Academic Medical Center with Established Restrictions)

Site A’s 116 modifications reflected high activity intensity but strong anticipatory planning (3.5:1 planned:unplanned ratio). There was a focus on training/education (n=30), reflecting the distinctive demands of an academic medical center with rotating resident/trainee populations and the hospital’s hybrid ICU model (combining dedicated intensivists and physicians with wider hospital responsibilities) that necessitated a wider range of physician engagement strategies.

Existing restriction infrastructure (prior carbapenem, ceftriaxone, and vancomycin restrictions) enabled leverage of established stewardship relationships and pre-existing familiarity with restrictions. FQ restriction represented “new additions to our restricted list” and enabled of technical integration with minimal change infrastructure modifications (n=2). Different stakeholder engagement was required because FQ prescribing patterns differed from previously restricted antibiotics. Surgical specialties, not heavily involved in prior restrictions, required tailored and directed engagement for FQ (commonly used for prophylaxis and empiric coverage).

#### Site B (Rural Community Hospital)

Site B generated the highest modification count (174) despite smallest organizational size, with rural context driving extensive adaptation needs. The 2.3:1 planned:unplanned ratio suggested anticipatory planning and reactive adjustment. Adaptation strategies showed the highest unplanned/reactive modifications (n=20), revealing unanticipated contextual demands despite forethought.

Organizational challenges inherent to rural community hospital settings shaped this implementation. Resource constraints included limited IT personnel (1 person on full-time effort for entire hospital) which created extended EHR build timelines (12+ months vs. 6 months at academic sites). The implementation team was required to modify stakeholder engagement extensively (33 stakeholder relationship modifications, exceeding all other sites), to maintain momentum during extended timelines.

Site B’s rural workforce characteristics (smaller staff, fewer specialists) created both facilitators and obstacles. Research naivety and limited experience with complex research interventions heightened sensitivity to implementation burden perceptions. The research team invested heavily in one-on-one conversations, adapting the intervention to existing workforce and workflow constraints, and ensuring stakeholder input shaped decisions.

COVID-19 caused the most significant disruptions in the study at Site B, suspending implementation for six months during pandemic peak when hospital operated at surge capacity. Re-engagement after suspension required rebuilding relationships and repeating educational activities, demonstrating the vulnerability of rural hospitals and implementation momentum to systemic disruptions.

#### Site C (Community Hospital with Established Stewardship)

Site C generated fewest modifications (94) but had the lowest planned:unplanned ratio (1.6:1. While this ratio indicates less anticipatory planning and is suggestive of lower organizational readiness, context matters for interpretation.

Site C’s established ICU workflow featured robust pharmacist-physician co-location and collaboration, integrated data systems, and strong quality improvement culture. These strengths facilitated rapid initiation, uptake and acceptability of FIRST in the ICU. FQ restriction represented “simply another layer” (quote from site implementation coordinator) rather than a radical change. Site C’s decision to implement FQ restriction hospital-wide (vs. ICU-only) represents a major proactive adaptation to prevent patient transfer workflow issues. however created scale-related challenges that could not be anticipated. They required an alternate authorization pathway (direct communication with antibiotic stewardship team, expanded authorization personnel) not outlined in the original implementation strategies.

Site C’s implementation approach was more iterative and feedback-responsive than at other sites. Use of evaluative strategies dominated Site C’s modifications (n=23), reflecting commitment to data-driven refinement of the implementation. The community hospital setting, with stable workforce no rotating trainees, enabled a focus on continuous improvement and deepened interprofessional trust between pharmacist-implementers and intensivists.

#### Site D (Academic Medical Center with Limited BPA Culture)

Site D’s 103 modifications and 1.8:1 planned:unplanned ratio reflected intermediate organizational preparedness. Their implementation was impacted by a recent Epic transition (18 months prior to FIRST) which, created IT resource competition, with multiple concurrent EHR optimization projects. Site D also had limited pre-existing BPA deployment, with a culture of privileging physician decision-making autonomy. These factors created unexpected contextual barriers, requiring technical implementation support alongside cultural change management. The relatively high proportion of unplanned training modifications (10) reflected an iterative approach to educating providers unused to clinical decision support restrictions. The dominance of evaluative strategies (predominantly planned at 15 vs. 6 unplanned) affirmed Site D’s data-driven implementation culture. The 1.8:1 planned-to-unplanned ratio and the “high touch” implementation approach to obtain senior physicians study buy-in demonstrate that organizational cultures vary, and even well-resourced academic centers may require intensive individualized engagement and ongoing adaptation when implementing novel interventions that are inconsistent with pre-existing site culture.

### 3.4 Planned vs. Unplanned Modifications Dynamics

Analysis of planned vs. unplanned modifications revealed three distinct modification triggers: workforce and prescriber dynamics, technological challenges, and external shocks.

Workforce demands generated the highest number of combined planned (n=60) and unplanned (n=28) modifications (ERIC strategy cluster “Train and educate stakeholders”) at all sites, generating a planned: unplanned ratio of 2.1:1. Resident rotations requiring repeated training (Sites A, D) and staff changes necessitated re-orientation about the intervention (Site B). Academic centers undergo predictable workforce cycles (resident rotations) that required strategy modifications in training content, timing, and reinforcement strategies. The planned:uplanned ratio indicates these modifications benefited from anticipatory planning. Contextual differences in technological elements necessitated both planned (n=46) and unplanned (n=40) modifications across all sites (ERIC strategy cluster “Adapt and tailor to context”). EHR customizations required workflow redesign and finessing of the components of the alert. Awareness of potential alert fatigue prompted alert refinement (Site C, D).

Unintended alert triggers (e.g., alerting for antibiotic orders that shouldn’t have triggered, not alerting when they should), and integration issues with other clinical decision support tools were discovered during trial runs (Sites A, C). Sites with recent EHR transitions or implementations (Sites B, D) experienced more technology-related unplanned modifications. These modifications generated a planned: unplanned ratio of 1.2:1 indicating a balance of anticipatory and reactive modifications.

External shocks, particularly COVID-19 pandemic impacts, generated major unplanned modifications. Site B’s six-month suspension represented the most severe disruption. Site A adapted by virtualizing grand rounds and educational sessions. Site C extended pre-implementation by one month to accommodate pandemic protocol prioritization. Site D experienced IT staff shortages as technical personnel were reassigned to pandemic-related systems needs.

The relationship between organizational experience and planned:unplanned ratios suggested that implementation infrastructure and prior experience with analogous interventions enhanced anticipatory planning capacity. Site A’s 3.1:1 ratio reflected extensive prior restriction experience enabling proactive strategy specification. Site C’s 1.6:1 ratio, despite established stewardship, reflected limited experience with hospital-wide EHR-based restrictions, revealing that stewardship maturity alone does not guarantee comprehensive anticipatory planning.

### 3.5 Phase-based timing of modifications

#### Pre-implementation

(pre-BPA launch), implementation modifications emphasized stakeholder development (train and educate stakeholders 21.3%, n=19) and consumer engagement (“Develop stakeholder interrelationships” and “Engage consumers” both at 16.9%, n=15), with a heavy focus on developing the pre-launch educational infrastructure for study participants (i.e. ICU prescribers). BPA customization, workflow plan integration, and unit-specific adaptations also predominated at this time (adapt and tailor to context 13.5%, n=12). Baseline data collection about site contexts, study monitoring system setup and pilot-testing of the BPA protocols also occurred (use of evaluative and iterative strategies 12.4%, n=11).

#### Early implementation

(<4 months post-BPA launch) showed total modifications were skewed by the continuous education burden of rotating trainees at academic Sites A and D, new rotations having entered during this period (train and educate stakeholders 38.5%, n=10). There was a shift to evaluative strategies (30.8%, n=8 vs. 12.4% pre-implementation) as data-driven process-refinement increased in response to early implementation challenges. Workflow adjustments, BPA firing refinements, and unit specific troubleshooting also occurred within this time-frame (adapt and tailor to context, 15.4%, n=4), especially at Site B.

#### Late implementation

(>4 months post-BPA launch) showed no implementation strategy modifications. This period also coincided with loss of contact with Site B, as the pandemic related impact interrupted research related activities at this site, and is subsequently a less accurate assessment of on-site implementation challenges and modifications for this site from this time onwards.

### 3.6 FRAME-IS Modification Characteristics

FRAME-IS module analysis revealed modification characteristics across dimensions:

Modification types (FRAME-IS Module 2): Content modifications (41.8% of total) dominated, reflecting changes to implementation strategy substance and delivery. Context modifications (25.1%) adapted strategies to local settings and infrastructure. Training modifications (17.4%) refined educational approaches. Evaluation modifications (15.7%) enhanced monitoring and iterative improvement processes.

Modification nature (Module 3): All sites maintained core EBP fidelity—the fundamental restriction mechanism remained unchanged (EHR alert requiring authorization for non-approved indications). Modifications targeted implementation processes: education timing, stakeholder engagement intensity, feedback mechanisms, workflow integration details. Fidelity-consistent adaptation characterized all sites.

Modification rationales (Module 4): Modifications operated primarily at organizational (40%) and implementer levels (35%), with fewer clinician-level (20%) or patient-level modifications (5%). Organizational-level rationales emphasized workflow efficiency, resource optimization, and integration with existing processes. Implementer-level rationales focused on enhancing implementation team capacity, improving stakeholder communication, and strengthening data systems. Clinician-level modifications addressed prescriber concerns, workflow impacts, and decision support usability.

Modification timing (Module 5): Pre-implementation phase generated 62% of planned modifications, focusing on stakeholder engagement, education development, and infrastructure building. Implementation phase (first 4 months post-launch) generated 71% of unplanned modifications, primarily addressing workflow issues, technical challenges, and resistance behaviors. Late implementation/maintenance phase (5-12 months post-launch) generated predominantly planned modifications for sustainability (ongoing training schedules, routine monitoring, institutionalization activities).

Decision-making (Module 6): Program leaders and implementation strategy experts collaboratively made >80% of modification decisions, with implementers initiating the majority. Site-initiated modifications substantially exceeded researcher-initiated modifications, demonstrating local adaptation authority and rapid decision-making capacity. Collaborative decision-making involved multidisciplinary teams, with modifications rarely made unilaterally.

Modification spread (Module 7): Unit-wide spread characterized infrastructure and policy modifications. Clinic/unit level spread also dominated workflow and process modifications (87%). Organization-wide spread (4.8%) reflected both study design and implementation approach. Individual level modifications (2%) targeted specific high-priority stakeholders or problematic workflows.

## 4.0 Discussion

### 4.1 Key Findings Summary

This study demonstrates that integrating ERIC and FRAME-IS frameworks enables comprehensive documentation and analysis of implementation strategy modifications across heterogeneous healthcare settings. Key findings include: (1) Planned modifications (68%) substantially outnumbered unplanned modifications (32%), but this ratio varied appreciably across sites (1.6:1 to 3.5:1), (2) Three ERIC clusters dominated modifications: stakeholder engagement, training/education, and evaluative strategies, (3) Rural hospitals and academic training centers exhibited distinct modification patterns related to contextual characteristics, (4) Prior experience with analogous interventions enhanced anticipatory planning capacity, and (5) All modifications maintained core EBP fidelity while adapting implementation processes.

### 4.2 Integration of Implementation Science Frameworks

ERIC provided standardized language for strategy identification and categorization, enabling systematic comparison across sites and time periods while FRAME-IS structured modification documentation. The resultant 80-modification range (94-174 total modifications) across sites demonstrated that organizational context shapes both strategy selection and modification intensity, an insight invisible when using either framework alone, highlighting the importance of accounting for organizational differences.

The planned vs. unplanned categorization, while subjective, provided a lens for analyzing implementation preparedness. Stirman et al (2019) and Miller et al (2021) suggest that planned-to-unplanned ratios may indicate pre-implementation assessment quality, with higher ratios showing comprehensive contextual assessment that enable anticipatory strategy selection. Similarly, we found higher planned:unplanned ratios in this study indicated robust anticipatory planning, strong stakeholder relationships that enabled proactive problem-solving, and organizational learning from previous implementations. Stirman et al. (2019) also suggest that lower ratios indicate reactive implementation requiring continuous adjustment, potentially reflecting limited implementation infrastructure or inadequate pre-implementation assessment.(13, 14) However, Site C’s implementation approach was evaluation-driven and iterative, rather than due to planning deficiencies, indicating that the planned-to-unplanned ratio was influenced as well by implementation philosophy at each site. The planned-to-unplanned distinction, when related to site heterogeneity, revealed which implementation domains benefit from anticipatory planning and which require reactive modification.

Our finding that planned modifications substantially outnumbered unplanned modifications (n=330 vs. 157 at all sites) challenges assumptions that implementation flexibility is primarily or necessarily reactive.

### 4.3 Organizational Context and Modification Patterns

Rural hospital characteristics drove specific modifications: extended timelines due to limited resources, intensive relationship-building due to smaller workforce and closer interpersonal dynamics, and creative problem-solving to overcome resource constraints. The presence of 24/7 antibiotic stewardship pharmacist support at Site B, while uncommon in rural settings, was accompanied by an existing workflow that required modification of the originating BPA to accommodate their site constraints.

Academic medical centers with rotating trainees faced perpetual education challenges. Resident rotations created predictable but nonetheless challenging training cycles. The distinction between Sites A and D highlighted how institutional BPA culture matters: Site A’s established alert familiarity enabled focusing on FQ-specific content, while Site D’s limited BPA approach to avoid “alert fatigue” and privilege physician autonomy required foundational education about clinical decision support concepts alongside FQ-specific training.

Community hospitals with established stewardship infrastructure could leverage existing relationships and systems but still required substantial modification when scaling interventions (ICU to hospital-wide) or changing restriction targets. Prior stewardship experience facilitated stakeholder engagement and workflow integration but did not eliminate modification needs, demonstrating that experience with one intervention type does not fully transfer to novel interventions.

Greenhalgh and colleagues classically characterize a site’s “absorptive capacity for new knowledge” as a critical organizational characteristic influencing innovation adoption.(24) We found a nuanced relationship between organizational absorptive capacity and total modification requirements. Sites having a greater organizational absorptive capacity due to a research-friendly and high-resource setting (i.e., Sites A and D) did not necessarily demonstrate lower required modifications. Structural features (resident rotations, open ICU models) can instead drive modification needs independently of organizational capacity. Conversely, despite being a community hospital with lower resources, Site C’s low modification count (n=94) demonstrates that mature stewardship culture with strong interprofessional bonds can compensate for resource limitations.

In keeping with Proctor and colleagues’(6) recommendation that implementation strategy temporality also be described, our analysis grouped ERIC clusters by the phases of implementation: pre-intervention, BPA-innovation launch, and sustainment. Pre-intervention (pre-BPA launch), strategy clusters were focused on increasing intervention acceptability and uptake, with the highest frequencies being: (1) train and educate stakeholders, and (2) develop stakeholder interrelationships and engage consumers, and (3) adapt and tailor to context. All strategies focused on capacity building through training and providing materials to increase provider skills, implementation process support in which direct help for planning and execution were provided, and integration strategies that tailored the implementation to the site workflows. Our results highlight the importance of increasing the “agreeability” of the strategy to participants, highly consistent with existing literature on acceptability.(25)

During the early implementation phase (<4 months post-BPA launch), sites were undergoing a process-driven refinement of their implementation strategies to meet the needs of site contexts and workflows. Frequency of ERIC clusters in this period shifted to evaluative and adaptive. These strategies were also aimed at increasing the intervention penetration and sustainability. The educational component of decreased in importance, except at Site A due to its continuous educational requirements.

There were few strategy modifications in the late implementation phase (4 and more months post-BPA launch), suggesting that the pre-and early-implementation periods are the most significant for implementing EHR-based, best practice alert-type interventions like the FIRST trial.

### 4.4 Implications for Implementation Science Theory

Our findings align with and extend several implementation science theories. The Consolidated Framework for Implementation Research (CFIR) emphasizes inner setting characteristics shaping implementation; our findings specify how characteristics like organizational size, academic status, and prior experience translate into concrete modification needs. Planned vs. unplanned modification tracking operationalizes concepts of continuous adaptation that align with other frameworks, such as the Dynamic Sustainability Framework.(2) Absorptive capacity theory gains empirical support through demonstrated relationships between prior experience and modification patterns.

This study advances understanding of implementation strategy adaptability. While maintaining core intervention fidelity, substantial implementation process modification was not just acceptable but necessary for contextually appropriate implementation. This challenges rigid protocol adherence models and supports flexible, learning-oriented implementation approaches.

### 4.5 Practical Recommendations for Implementation

For rural hospitals: Budget extended pre-implementation phases (12+ months), front-load stakeholder engagement extensively (30+ stakeholder activities), build substantial adaptation flexibility into protocols, plan for limited IT resources with external technical support, establish 24/7 coverage through creative staffing solutions, create contingency plans differentiating essential vs. deferrable activities, and expect modifications to continue after intervention launch.

For academic medical centers with rotating trainees: Design perpetual education systems with systematic orientation content for every rotation cycle, tailor approaches by specialty (direct faculty engagement for surgical specialties, resident-focused education for medicine), leverage existing educational infrastructure (grand rounds, teaching conferences, handshake rounds), build automated data extraction systems, plan monthly re-education, and focus on establishing sustainable processes rather than one-time training events.

For community hospitals with established stewardship: Leverage existing relationships and pharmacist-physician collaboration, strategically consider intervention scope (hospital-wide vs. ICU-only) based on capacity and transfer patterns, engage IT teams early given personnel availability constraints, establish intensive post-implementation monitoring with data-driven refinement, prepare alternative workflows if private/contracted groups decline participation, front-load consumer engagement and stakeholder relationship development, and expect modifications emphasizing evaluative strategies and infrastructure adjustments.

For academic centers new to clinical decision support: Invest in cultural change management addressing BPA resistance through senior physician champions, implement “high-touch” engagement with individualized meetings and practice pattern data, build robust data monitoring with automated reporting and monthly feedback, plan for iterative training adjustments based on initial experiences, leverage existing research infrastructure (IRB, data services, coordinators), and avoid assuming academic status eliminates resistance—prescriber autonomy culture may create significant barriers requiring proactive engagement strategies.

### 4.6 Limitations

We selected sites for high variability to increase comparison for this analysis, and while the methodology remains transferrable, the small sample (n=4 sites) limits generalizability of site-level patterns. Future research with larger sample sites could establish more robust associations between organizational characteristics and modification patterns. Our data collection and analysis were also resource-intensive, requiring multi-source documentation and substantial analytical time for coding, consensus discussions, and cross-site synthesis. Not all implementation studies may have resources for this level of effort. More explicit advance documentation of planned strategies and their modifications may also be beneficial for future analyses. While all sites achieved successful implementation (as assessed by adoption, reach, and acceptability), we cannot determine which specific modifications were most critical to success versus which were less impactful. Detailed fidelity assessment beyond what is captured in the FRAME-IS modules was also beyond the scope of this paper.

#### COVID-19 Differential Impact

The COVID-19 pandemic’s differential impact across sites illuminates organizational resilience patterns. Site B**’**s severe disruption (6-month research suspension) versus Site A**’**s minimal impact (virtual grand rounds) reflects rural-urban resource disparities and research infrastructure robustness. Sites with established stewardship cultures (Site C, Site D) demonstrated greater pandemic resilience, suggesting that mature quality improvement infrastructure provides buffering capacity during systemic crises. Despite pandemic-related challenges, all sites successfully implemented the intervention, though timelines varied considerably (Site B experienced the most substantial delay).

## 5.0 Conclusion

This four-site case study analysis demonstrates that successful restrictive stewardship intervention implementation requires contextual awareness, implementation support intensity calibration, and an awareness of strategies specific to the implementation phases of each site. The modification range (94-174 total modifications) across sites implementing an identical core intervention underscores that “one size fits all” implementation approaches fail to account for critical organizational differences.

Four key insights emerge. Firstly, organizational context (readiness, relational collaboration between implementation champions and the implementers, leadership support, site implementation coordinator authority and support, resource limitations, educational context and more) determines which strategies are needed, when they will be needed, and how intensively they must be deployed. Academic centers may require disproportionate training investment due to rotating trainee populations; rural hospitals may require extensive adaptation capacity and personnel support due to resource constraints; diverse health systems may require cultural change management when introducing novel interventions.

Secondly, planned:unplanned modification ratios provide novel indicators of organizational implementation readiness and anticipatory planning capacity. Higher ratios (3:1) characterized organizations with prior implementation experience, established infrastructure, and strong stakeholder relationships, suggesting robust capacity for proactive strategy adaptation. Lower ratios (1.6:1) characterized organizations implementing novel intervention types or lacking implementation infrastructure, indicating reactive implementation requiring continuous adjustment and potentially greater support needs. Similarly, high modification counts do not signal implementation failure; rather, they may reflect intentional iterative refinement or proactive anticipation of contextual barriers. The distinction lies between planned adaptations demonstrating foresight and unplanned modifications indicating unanticipated barriers, an insight best derived from qualitative assessments of site contexts.

Thirdly, certain implementation domains universally benefit from front-loaded investment (infrastructure, stakeholder engagement) while others inherently require ongoing reactive modifications (training in academic centers, evaluation in data-mature organizations). Implementation strategy plans should allocate effort in a context-specific manner, rather than distributing effort uniformly across all domains.

Fourthly, implementation content requirements vary throughout the implementation process, and in accordance with site characteristics. Anticipating and designing tailored implementation strategies informed by these phases can reduce the implementation burden accordingly.

In conclusion, the integrated ERIC-FRAME-IS approach with explicit planned/unplanned coding provides a systematic method for tailoring implementation strategies to organizational context. Future research should: (1) examine whether planned:unplanned ratios predict implementation outcomes, (2) investigate optimal modification timing and sequencing, (3) explore modification patterns across diverse intervention types and healthcare settings, (4) develop tools for organizational implementation readiness assessment predicting likely modification needs, and (5) study sustainability modifications during post-implementation phases.

## Data Availability

No datasets were generated or analysed during the current study. All relevant data from this study will be made available upon study completion.

## Acknowledgements

The authors gratefully acknowledge the dedication and expertise of our site implementation coordinators and champions whose commitment to improving antimicrobial stewardship and patient safety made this work possible. We thank the infectious disease physicians, pharmacists, stewardship program leaders, information technologists, nursing staff, and clinical leadership at all participating hospitals. Special recognition goes to our site implementation coordinators who maintained detailed implementation documentation and generously shared their experiences throughout this study. We acknowledge the patients whose care motivated our collective commitment to reducing healthcare-associated infections.

## Ethical approval and informed consent statement

The study’s protocol was approved by the University of Wisconsin-Madison’s Health Sciences Institutional Review Board and that of the other participating sites.

